# Mapping the Regional Distribution of Molecular Tumor Board Patients and Identifying White Spots in our Catchment Area – a Joint Approach of the CCC WERA Alliance

**DOI:** 10.1101/2022.05.15.22275106

**Authors:** Florian Lüke, Florian Haller, Kirsten Utpatel, Markus Krebs, Norbert Meidenbauer, Alexander Scheiter, Silvia Spörl, Daniel Heudobler, Felix Keil, Christoph Schubart, Lars Tögel, Sabine Einhell, Wolfgang Dietmaier, Ralf Huss, Sebastian Dintner, Sebastian Sommer, Frank Jordan, Maria-Elisabeth Goebeler, Michaela Metz, Diana Haake, Mithun Scheytt, Elena Gerhard-Hartmann, Katja Maurus, Stephanie Brändlein, Andreas Rosenwald, Arndt Hartmann, Bruno Märkl, Hermann Einsele, Andreas Mackensen, Wolfgang Herr, Volker Kunzmann, Ralf Bargou, Matthias W. Beckmann, Tobias Pukrop, Martin Trepel, Matthias Evert, Rainer Claus, Alexander Kerscher

**Affiliations:** Department of Internal Medicine III, University Hospital Regensburg, Hematology and Oncology, Regensburg, Germany; Comprehensive Cancer Center Ostbayern, Regensburg, Germany; Division of Personalized Tumor Therapy, Fraunhofer Institute for Toxicology and Experimental Medicine, Regensburg, Germany; Institute of Pathology, Friedrich-Alexander University Erlangen-Nuremberg, University Hospital Erlangen, Erlangen, Germany; Comprehensive Cancer Center Erlangen-EMN, Erlangen, Germany; Institute of Pathology, University of Regensburg, Regensburg, Germany; Comprehensive Cancer Center Mainfranken, University Hospital Würzburg, Würzburg, Germany; Department of Urology and Pediatric Urology, University Hospital Würzburg, Würzburg, Germany; Department of Medicine V, Hematology and Oncology, University Hospital Erlangen, Erlangen, Germany; Bavarian Center for Cancer Research / BZKF, Bavaria, Germany; Institute of Pathology and Molecular Diagnostics, Medical Faculty, University of Augsburg, Augsburg, Germany; Comprehensive Cancer Center Augsburg, Augsburg, Germany; Department of Hematology and Clinical Oncology, Medical Faculty, University of Augsburg, Augsburg, Germany; Department of Internal Medicine II, University Hospital Würzburg, Würzburg, Germany; Institute of Pathology, University of Würzburg, Würzburg, Germany; Department of Gynecology and Obstetrics, University Hospital Erlangen, Erlangen, Germany

**Keywords:** Precision Oncology, MTB, Patient Access, Cancer Care, Outreach, Real world data, Outcomes Research

## Abstract

**Background:** Molecular Tumor Boards (MTBs) are crucial instruments for discussing and allocating targeted therapies to suitable cancer patients based on genetic findings. Currently, limited evidence is available regarding the regional impact and the outreach component of MTBs.

**Methods:** We analyzed MTB patient data from four neighboring Bavarian tertiary care oncology centers in Würzburg, Erlangen, Regensburg, and Augsburg, together constituting the WERA Alliance. Absolute patient numbers and regional distribution across the WERA-wide catchment area were weighted with local population densities.

**Results:** Highest MTB patient numbers were found close to the four cancer centers. However, peaks in absolute patient numbers were also detected in more distant and rural areas. Moreover, weighting absolute numbers with local population density allowed us to identify regions within our catchment area relatively underrepresented in WERA MTBs.

**Conclusion:** Investigating patient data from four neighboring cancer centers, we comprehensively assessed the regional impact of our MTBs. The results confirmed the success of existing collaborative structures with our regional partners. Additionally, our results help identifying potential white spots in precision oncology and establishing a joint WERA-wide outreach strategy.

## INTRODUCTION

Precision oncology has made immense progress in delivering novel therapies guided by molecular biomarkers to patients suffering from cancer. This was made possible by the rapid advancements in molecular diagnostics. While generating mutational profiles has become feasible and readily available, interpretation of mutational profiles and integration of molecular and clinical data for therapeutic recommendations is still a challenge. Molecular Tumor Boards (MTBs) usually located at tertiary care oncology centers have therefore become groundbreaking and indispensable institutions for attributing specific drugs to suitable patients based on individual tumor biology. Within MTBs, potential therapeutic strategies are discussed by clinicians, pathologists, and researchers such as molecular pathologists and human geneticists.

Despite its promises, there are concerns that precision oncology programs could exacerbate health disparities within societies by excluding patients in underserved regions and patients from underserved communities from these promising treatment options [1]–[4]. When setting up the “National Decade against Cancer” in 2020, the German Federal Ministry of Education and Research therefore decided to put a special emphasis on providing equal access to precision oncology for all patients in Germany [5]. In line with this goal, the four Bavarian tertiary care oncology centers in Würzburg (W), Erlangen (E), Regensburg (R), and Augsburg (A) founded the WERA Alliance to provide equal access to precision oncology for all patients from its mainly non-metropolitan/rural catchment area. Figure 1 outlines the catchment area of the WERA Alliance with its regional partner hospitals which covers a large part of the Free State of Bavaria.

**Figure 1:**
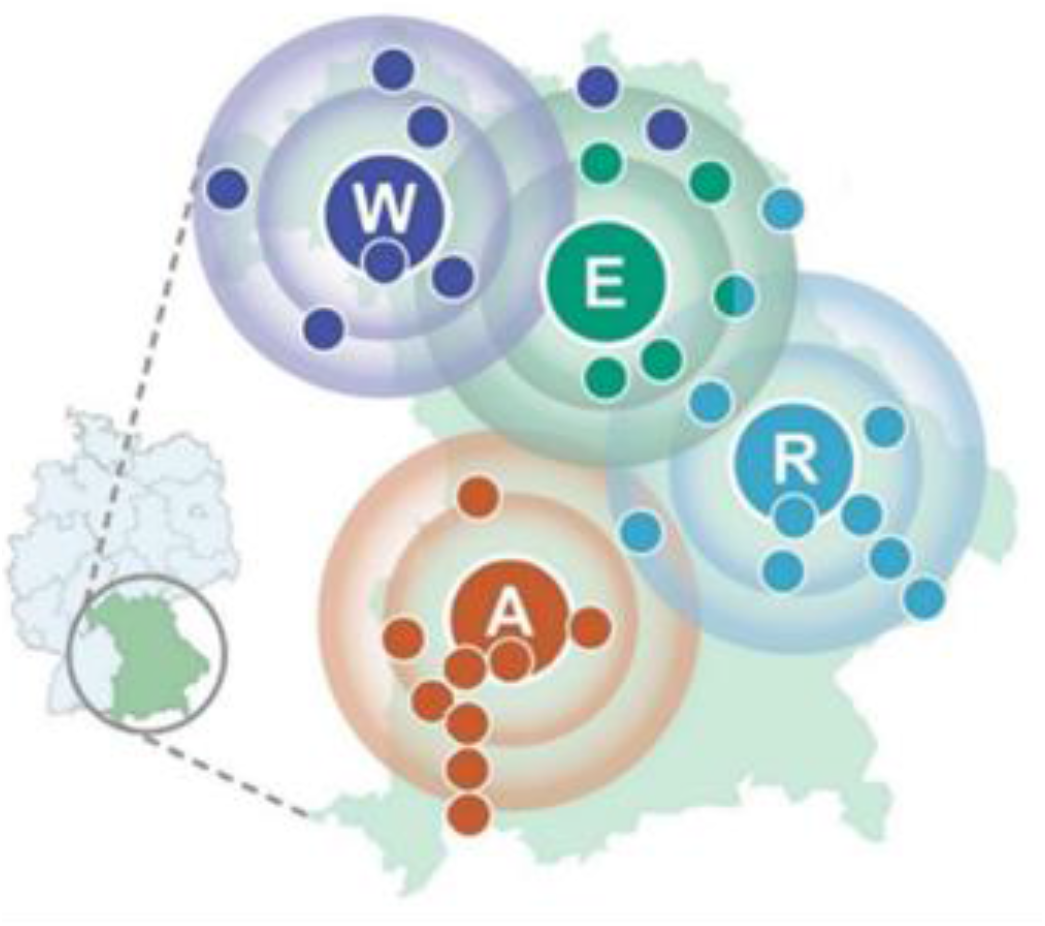
Regional catchment area of the WERA cancer center alliance – containing Würzburg (W), Erlangen (E), Regensburg (R), and Augsburg (A) as regional hubs with cooperating regional hospitals (colored smaller dots) – plotted on a map of the Free State of Bavaria in Germany.

As representatives of the MTBs at all four cancer centers, we collectively measured the regional impact of our boards by mapping the physical addresses of our patients discussed between 2020 and 2021. Absolute patient numbers were weighted with local population densities in order to identify regions of our joint catchment area, which are underrepresented in WERA MTBs.

## MATERIALS AND METHODS

For this retrospective analysis, we collected postal codes from physical addresses (at time of board discussion) of all MTB patients at the university hospitals of Würzburg, Erlangen, Regensburg, and Augsburg in the years 2020 and 2021. Data were provided by local tumor registries of each university hospital and, if necessary, local hospital information systems. This study was performed in accordance with local GDPR and the Bavarian Hospital Act (“Bayerisches Krankenhausgesetz”).

German population data for respective postal code areas were downloaded from an open-source database (https://www.suche-postleitzahl.org/downloads) – combining German postal code information provided by OpenStreetMap (https://www.openstreetmap.org) with population data from German statistical offices within the “Zensus 2011” initiative (https://www.zensus2011.de). For weighting patient numbers with local population densities, we determined the number of MTB patients per 100,000 inhabitants (termed: local patient representation). Data merging and curation were performed with Access (Microsoft®, Redmond, WA), calculations and illustration (including mapping) were performed with Excel (Microsoft®, Redmond, WA).

## RESULTS

### Characterizing MTB patients from Würzburg, Erlangen, Regensburg, and Augsburg

For the years 2020 and 2021, we examined the regional provenance of patients discussed in MTBs at the university hospitals of Würzburg, Erlangen, Regensburg, and Augsburg. Detailed patient characteristics for each WERA MTB are summarized in Table 1.

**Table 1:** Characteristics of our joint study cohort for each WERA MTB. Regarding local patient representation, absolute patient numbers (n) were weighted with local population (n/100,000 inhabitants). IQR: Interquartile range.

|  | Patient numbers (n) | Patient numbers per postal code area |  |  |  |
| --- | --- | --- | --- | --- | --- |
|  |  | Max. local patient numbers (n) | Max. local patient representation (n/100,000 inhabitants) | Median local patient numbers (n); [IQR] | Median local patient representation (n/100,000 inhabitants); [IQR] |
| Würzburg | 385 | 8 | 194.17 | 1 [1] | 24.35 [32.11] |
| Erlangen | 521 | 19 | 294.99 | 1 [1] | 27.39 [32.76] |
| Regensburg | 217 | 7 | 111.23 | 1 [1] | 18.93 [24.21] |
| Augsburg | 251 | 11 | 251.57 | 1 [2] | 27.64 [32.83] |
| WERA Alliance | 1,374 |  |  |  |  |

In total, our study analyzed the regional origin of 1,374 MTB patients – with 385 patients from Würzburg and 521 from Erlangen. Regensburg and Augsburg contributed 217 and 251 MTB patients, respectively. We next calculated (absolute) patient numbers for each postal code area of the WERA outreach. Maximum local patient numbers per postal code area ranged from 7 (Regensburg) to 19 (Erlangen). After weighting patient numbers with local population per postal code area, maximum local patient representation ranged from 111.23 pts./100,000 inhabitants (Regensburg) to 294.99 pts./100,000 inhabitants (Erlangen). To further characterize and illustrate our current regional impact, we plotted absolute numbers of MTB patients from all four sites on a map of Southern Germany. Figure 2 illustrates our results.

**Figure 2:**
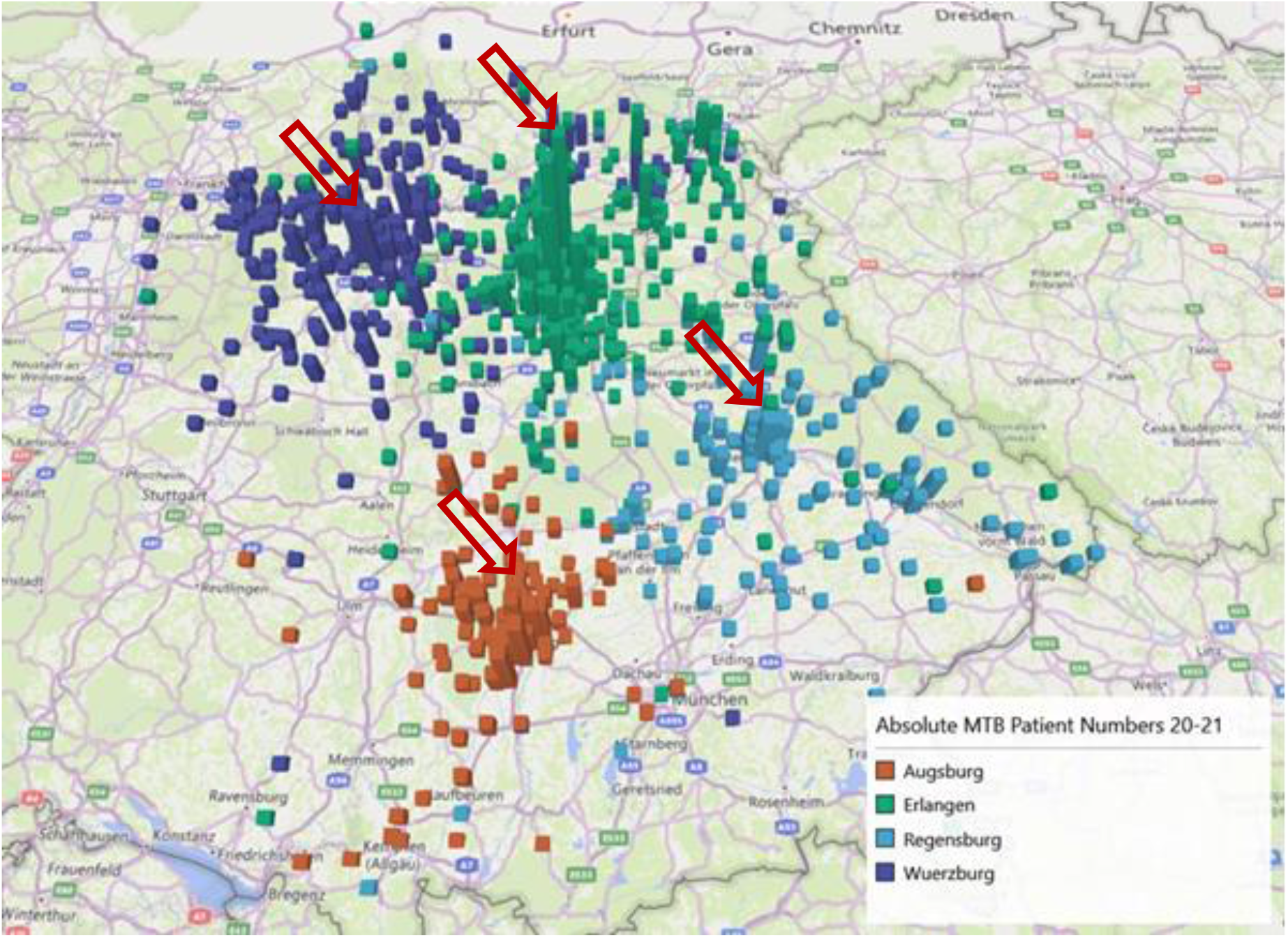
Absolute numbers of patients discussed in WERA MTBs in the years 2020 and 2021. Results are plotted on a map of the Free State of Bavaria and surrounding regions. Arrows mark peaks in absolute MTB patient numbers close to the four WERA university hospitals.

At first sight, WERA MTBs complemented each other well in terms of regional patient distribution – together already covering a substantial part of the joint catchment area shown in Figure 1. Of note, MTB patients did not exclusively live in the Free State of Bavaria, but also in the neighboring Federal States of Baden-Württemberg, Hesse, and Thuringia. Peaks in absolute patient numbers were seen for regions close to the four WERA university hospitals – as indicated by arrows in Figure 2.

However, we also identified clusters of patients beyond urban areas – for example, Kulmbach in the northeastern part and the region around Straubing, Deggendorf and Passau (partners of CCC Ostbayern) in the eastern part of Bavaria.

### Relative regional representation of cancer patients in WERA MTBs

To account for overrepresentation of urban areas with higher population densities and to allow a more differentiated view on our regional impact, we weighted absolute MTB patient numbers with local population density for each postal code area. This data transformation step highlighted existing networking structures of each WERA site by revealing “novel” peaks in rural areas and in the periphery of our catchment area (as indicated by arrows in Figure 3). Successful outreach activity was also reflected by similar measures of dispersion across all WERA sites regarding median local patient representation (Table 1).

**Figure 3:**
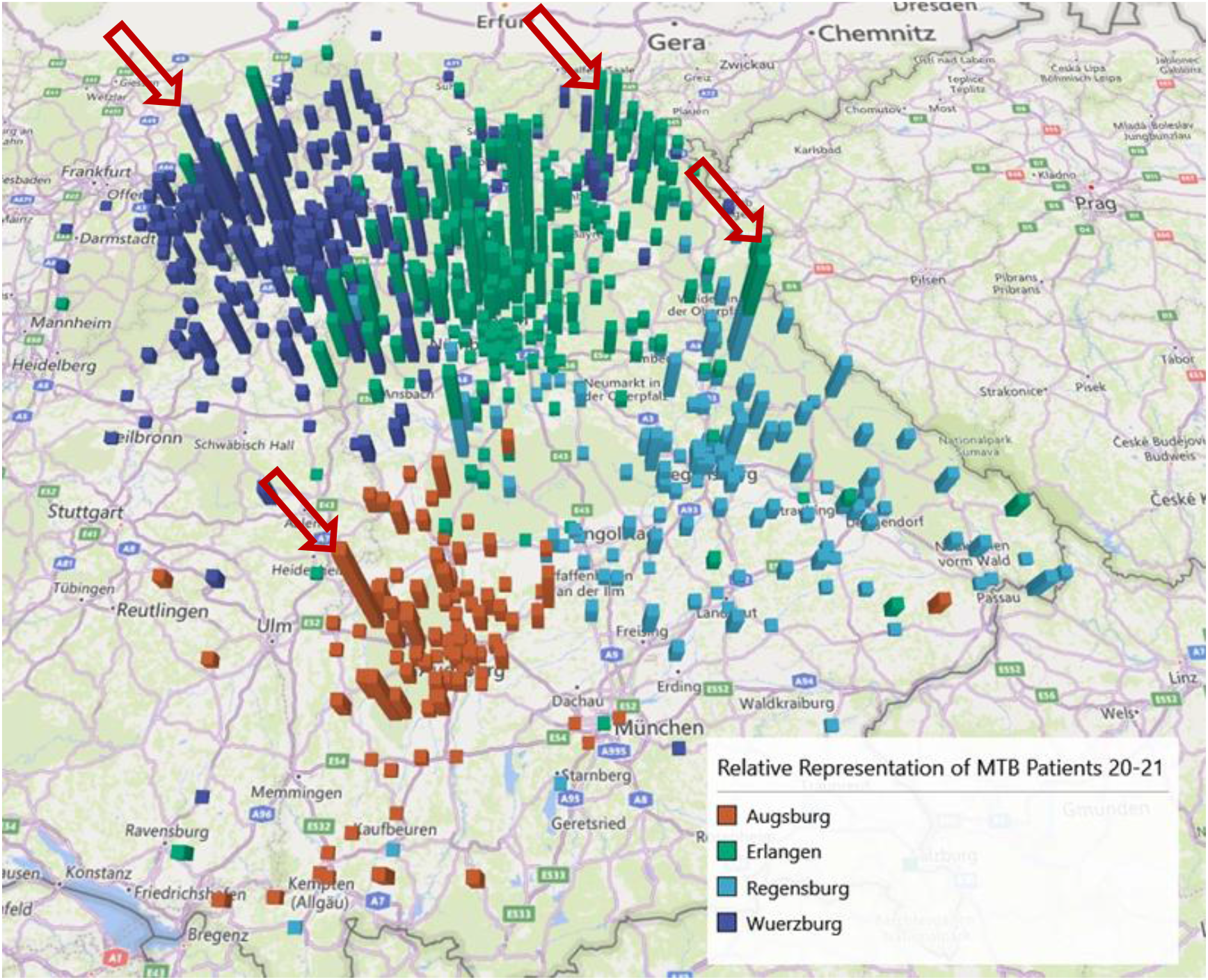
Local representation of patients discussed in WERA MTBs in the years 2020 and 2021. Absolute numbers were weighted with local population densities (MTB patients per 100,000 inhabitants). Arrows mark peaks in local representation reflecting successful network collaboration.

Changing diagrams from bar graphs to heat plots (Figure 4) allowed us to precisely locate postal code areas strongly represented in WERA MTBs during the recent two years – as well as areas, which were underrepresented at the same time. As shown in Figure 4A, postal code areas close to Aschaffenburg (No. 1) in the northwestern part of Bavaria displayed high counts in local patient representation. Moreover, WERA MTBs also discussed a high number of patients living in areas such as Bamberg (No. 2), Kulmbach (No. 3), and Rothenburg ob der Tauber / Bad Windsheim (No. 4). Regarding strongly represented areas close to the WERA cancer centers in Regensburg and Augsburg, we identified the rural area around Neunburg vorm Wald (No. 5) and the Günzburg / Burgau region (No. 6), respectively.

**Figure 4:**
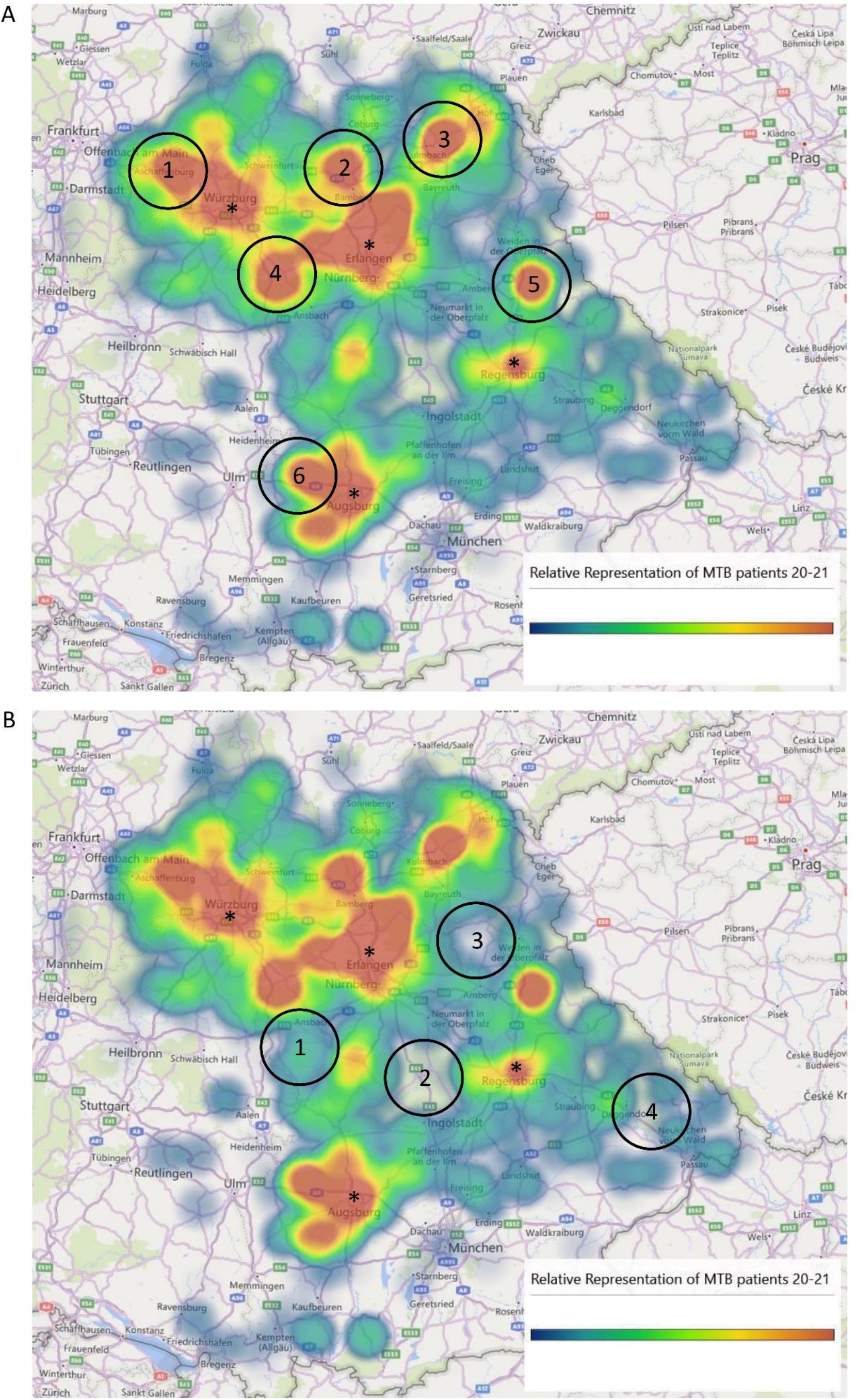
Heat plots illustrating the relative representation of MTB patients in 2020 and 2021 across the WERA catchment area (MTB patients per 100,000 inhabitants). (A) Indicated regions with high values reflect successful clinical collaboration and outreach activity. (B) Marked regions were underrepresented in WERA MTBs. (A, B) Asterisks mark the location of the WERA university hospitals.

As illustrated in Figure 4B, we also were able to delineate postal code areas, which were underrepresented in MTBs. This was the case for the region close to Ansbach (No. 1) and the rural area between Nuremberg and Ingolstadt (No. 2). Interestingly, area No. 3 contains the military training ground Grafenwöhr – basically representing an uninhabited region. The Grafenau region (No. 4) as well as neighboring regions close to the Czech border also emerged as white spots, possibly due to the fact that there are few practices specialized in hematology or oncology in these rural area in a certain radius.

## DISCUSSION

### Gaining insight through cooperation and joint data analysis

Various studies analyzed the MTB organizational structure as well as its impact on clinical decision-making and its benefit for cancer patients [6]–[9]. However, there is scarce evidence regarding the outreach component of MTBs located at tertiary care oncology centers. In our retrospective analysis, we therefore investigated the regional distribution of patients discussed in MTBs of the Bavarian university hospitals of Würzburg, Erlangen, Regensburg, and Augsburg – together constituting the WERA Alliance.

By jointly investigating our patient-centered care, we assessed our current impact on precision oncology within the WERA-wide catchment area. Regarding absolute patient numbers, highest peaks were seen for areas close to our four university hospitals. This result was not surprising, as it reflects high numbers of cancer patients primarily treated at our tertiary care centers as well as higher population densities in these metropolitan areas. However, we also found peaks in rural areas, which reflect existing and successful collaboration with regional health care providers. Altogether, mapping MTB patients from all four cancer centers confirmed the existing widespread regional impact of the WERA Alliance.

### Identifying potential white spots in precision oncology

To account for heterogenous population densities, relative representation of a certain postal code area within MTBs was defined as patients per 100,000 inhabitants. This approach even more highlighted established networking with regional partner hospitals and oncologists in private practices. These results underline that regional networks substantially increase treatment options for patients with cancer living in rural areas.

In contrast to rural areas strongly represented in WERA MTBs, we could also identify potential white spots within our catchment area – regions which were underrepresented in MTBs. In future, we need to have a thorough look at each of these white spots to specifically understand underlying causes for this statistical underrepresentation. In general, there could be a lack of information and awareness regarding the benefit of precision oncology programs with both, health care providers and patients. Another reason for potential white spots – demonstrated by the military training ground in Grafenwöhr – could be sparse overall population of certain regions. Moreover, white spots in our WERA-wide analysis might be covered by MTBs of different cancer centers. In our case, physicians might send their patients to one of the two MTBs performed at the university hospitals of Munich, which currently are not part of this analysis. As a consequence, each white spot candidate in our catchment area requires an in-depth analysis – above all to identify regions where a lack of information and awareness causes underrepresentation.

In our view, being able to locate these potentially underserved regions clearly shows the benefit of our joint approach, as no single-center analysis can address such a research question. These “negative results” will support WERA’s precision oncology policy by directing our outreach measures specifically towards underserved areas.

### Limitations and future directions

Our study has several limitations. Firstly, we use a simplistic model with the basic assumption of equally distributed cancer incidences across our catchment area. More specifically, we did not account for differences in cancer risk factors such as the age of the local population. However, given that MTBs cover all tumor entities, and the influence of certain risk factors is not equally distributed between cancer entities, we decided against stratifying for these risk factors. As already stated above, we cannot rule out that patients from underrepresented regions are sent to another tertiary care oncology center outside the WERA Alliance, especially in the periphery of our catchment area. Yet, a systematic bias appears improbable for the inner part of our catchment area constituted by our four neighboring centers. Last, we should state that patients discussed within MTBs clearly represent the “tip of the iceberg” in precision oncology, as many targeted therapies are also discussed and attributed within organ-specific tumor boards.

Due to these limitations, our study clearly has an exploratory and descriptive character. Yet, we are convinced that it is a further step to harmonize our outreach policy and to get a deeper understanding of what is needed to provide precision oncology programs for our rural catchment area. In the future, we will refine our analysis by considering local cancer incidences, MTB-specific distributions of cancer entities, and local demographic factors. Measures to improve our joint precision oncology program will include the integration of patient representatives and advocacy groups to raise awareness in the patient community. Moreover, information campaigns together with local healthcare providers and medical associations will provide valuable feedback on how to further improve accessibility in rural areas.

## Data Availability

All data produced in the present study are available upon reasonable request to the authors

## ACKNOWLEDGEMENTS

The authors thank Jörg Fuchs (University of Würzburg) for illustrating the outreach of the WERA Alliance (Figure 1).

